# Phylogeography and Resistome Epidemiology of Gram-Negative Bacteria in Africa: A Systematic Review and Genomic Meta-Analysis from a One-Health Perspective

**DOI:** 10.1101/2020.04.09.20059766

**Authors:** John Osei Sekyere, Melese Abate Reta

**Author notes:** **Address correspondence to:** Dr. John Osei Sekyere, Department of Medical Microbiology, School of Medicine, Faculty of Health Sciences, University of Pretoria, South Africa.; Tel. +27-*(0)12-319-2251.

## Abstract

**Objectives/Backgrounds:** Antibiotic resistance (ABR) remains a major threat to public health and infectious disease management globally. However, ABR ramifications in developing countries is worsened by limited molecular diagnostics, expensive therapeutics, inadequate skilled clinicians and scientists, and unsanitary environments.

**Methods:** Data on specimens, species, clones, resistance genes, mobile genetic elements, and diagnostics were extracted and analysed from English articles published between 2015 and December 2019. The genomes and resistomes of the various species, obtained from PATRIC and NCBI, were analysed phylogenetically using RAxML and annotated with Figtree. The phylogeography of resistant clones/clades was mapped manually.

**Results & conclusion:** Thirty species from 31 countries and 24 genera from 41 countries were respectively analysed from 146 articles and 3028 genomes. Genes mediating resistance to β-lactams (including *bla*_TEM-1_, *bla*_CTX-M_, *bla*_NDM_, *bla*_IMP_, *bla*_VIM_, *bla*_OXA-48/181_), fluoroquinolones (*oqxAB, qnrA/B/D/S, gyrA/B* and parCE mutations etc.), aminoglycosides (including *armA, rmtC/F*), sulphonamides (*sul-1/2/3*), trimethoprim (*dfrA*), tetracycline (*tet*(A/B/C/D/G/O/M/39)), colistin (*mcr-1*), phenicols (*cat*A/B, *cml*A), and fosfomycin (*fos*A) were mostly found in *Enterobacter spp*. and *K. pneumoniae*, and also in *Serratia marcescens, Escherichia coli, Salmonella enterica, Pseudomonas, Acinetobacter baumannii*, etc. on mostly IncF-type, IncX_3/4_, ColRNAI, and IncR plasmids, within *IntI*1 gene cassettes, insertion sequences and transposons. Clonal and multiclonal outbreaks and dissemination of resistance genes across species, countries and between humans, animals, plants and the environment were observed; *E. coli* ST103, *K. pneumoniae* ST101, *S. enterica* ST1/2 and *V. cholerae* ST69/515 were common strains. Most pathogens were of human origin and zoonotic transmissions were relatively limited. One Health studies in Africa are needed.

**Highlights/Significance:** Antibiotic resistance (AR) is one of the major public health threats and challenges to effective containment and treatment of infectious bacterial diseases worldwide. Herein, we used different methods to map out the geographical hotspots, sources and evolutionary epidemiology of AR. *Escherichia coli, Klebsiella pneumoniae, Salmonella enterica, Acinetobacter baumanii, Pseudomonas aeruginosa, Enterobacter spp*., *Neisseria meningitis/gonnorhoeae, Vibrio cholerae, Campylobacter jejuni* etc. were common pathogens shuttling AR genes. Transmission of same clones/strains across countries and between animals, humans, plants and the environment were observed. We recommend *Enterobacter spp*. or *K. pneumoniae* as better model/index organisms for AR surveillance.

## Introduction

Antibiotic resistance (ABR), particularly in Gram-Negative bacteria (GNB), is complicating infection management in Africa and the rest of the world as it restricts effective therapeutic options available to clinicians in human and veterinary medicine ^1–3^. Given the unsanitary environments common in developing countries as well as the limited healthcare and laboratory facilities, poor sewage management, high patient-to-physician ratios, little or no regulation on antibiotic usage etc., the escalation of ABR in developing countries (Africa) is precarious ^3–5^. Coupled with these is the limited funding available for molecular surveillance of ABR to map out the true burden of the problem ^2,4^. A recent global metagenomic survey, for instance, found that African countries had the highest ABR genes (ARGs) abundance, although in Animals, Asia was found to have more ABR hotspots than Africa ^2,5^.

Together, GNB cause or aggravate some of the most fatal and common infections and diseases known to mankind: sepsis, meningitis/meningococcemia, gonorrhoea, pneumonia, cystic fibrosis, urethritis, pelvic inflammatory disease, cholera, typhoid, whooping cough, diarrhoea etc^6–15^. It is worth noting that compared to Gram-positive bacteria, GNB has been implicated in the development of more resistance mechanisms, including resistance to last-resort antibiotics such as colistin (*mcr*), carbapenems (*bla*_NDM_, *bla*_VIM_, *bla*_KPC_, *bla*_IMP_, *bla*_GES_ etc.) and tigecycline (*tet(X)*) ^1,16,17^. Moreover, these ARGs are found in more diverse GNB species and genera, with substantial mortalities, morbidities and multi-drug resistance ^16–19^. Hence, GNB species such as *Pseudomonas aeruginosa, Acinetobacter baumannii*, and Enterobacteriaceae expressing resistance to carbapenems and extended-spectrum β-lactamases (ESBLs) are respectively classified as critical and high priority pathogens by the WHO 20,21.

The presence of these ARGs and species in human, environmental and animal specimens in Africa (and globally) are well documented, strengthening the call for further One Health molecular surveillance ^2,3,5,22^. Yet, a systematic review on GNB in Africa from a One Health perspective is lacking ^22,23^. This limits the comprehensive appreciation of the epidemiology of ARGs across the continent. Herein, we further undertake a detailed phylogenomic, phylogeographic and resistome analyses using publicly available genomes to augment the findings in the literature and describe the evolutionary epidemiology of ARGs in Africa.

## Methods

### Databases and Search Strategy

A comprehensive literature search was carried out on PubMed and/or ResearchGate, ScienceDirect and Web of Science electronic databases. English research articles published between January 2015 and December 2019 were retrieved and screened using the following search terms and/or phrases: “molecular epidemiology”, “gram-negative bacteria”; “mechanisms of resistance”; antimicrobial resistance genotypes”; “drug resistance”; “AMR genotypes”; “genetic diversity”; “clones”; “genotyping”; “antibiotic resistance gene”; “plasmid”; “mobile genetic elements”; “resistome”; “gene mutation”; “frequency of resistance gene mutation”; and Africa. The search terms used were further paired with each other or combined with the name of each African country, and searching strings were implemented using “OR” and “AND” Boolean operators.

### Inclusion and Exclusion Criteria

Articles addressing the molecular mechanisms (using PCR, microarray or WGS) of ABR in GNB and undertaking bacterial typing (MLST, PFGE, ERIC-PCR, WGS etc.) were included in this systematic review. Papers which addressed ABR using only disc diffusion, broth microdilution, E-test, Vitek, MicroScan etc. without a molecular (PCR/microarray/WGS) analyses of the underlying resistance mechanism and typing (clonality) of the isolates were excluded.

### Included data

This review was carried out using the Preferred Reporting Items for Systematic Reviews and Meta-Analyses (PRISMA) guidelines (Figure 1). The following data were extracted from the included articles: country; study year; specimen type and source/s; sample size; total isolates (isolation rate); total isolates for which antimicrobial sensitivity testing (AST) was determined; bacterial species; clone/MLST; antibiotic resistance gene (ARGs); mobile genetic elements (MGEs); antibiotic resistance phenotype; genotyping and diagnostic method(s)/techniques used (Tables S1-S3).

**Fig. 1:**
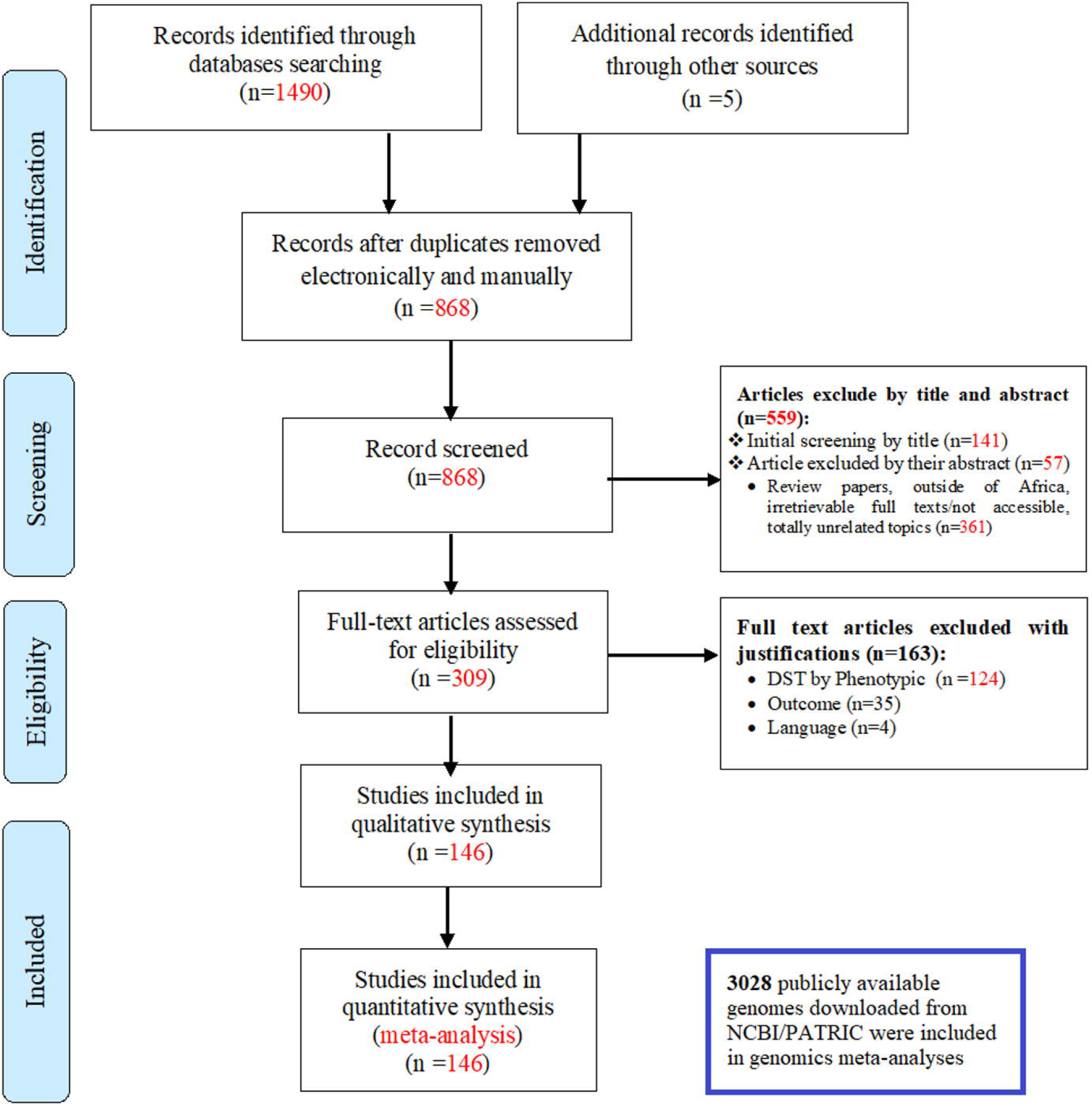
The PRISMA flow-diagram was used to summarise the literature search methodology, databases used, and the inclusion and exclusion criteria adopted in getting the final 146 manuscripts used in this qualitative and quantitative analyses. Besides the included literature, 3028 publicly available genomes were downloaded from PATRIC (https://www.patricbrc.org/)/NCBI’s Genbank and analysed to determine their resistomes and evolutionary phylogeography.

### Statistical and bio-informatic analyses

Microsoft Excel^®^ 365 was used to analyse the frequencies using raw data extracted from all included articles. To each GNB species, the resistance rates per antibiotic per country were calculated by dividing the total resistant isolates by the total isolates for which antibiotic sensitivity was determined. Frequencies of resistant species, clones, ARGs-MGEs associations and ABR rates were evaluated per animal, human and environmental sources per country (Tables S1-S3).

Genomes of 24 genera that were found in the included articles were downloaded from PATRIC (https://www.patricbrc.org) and used for phylogenetic analyses using RAxML’s maximum-likelihood method and annotated with Figtree (http://tree.bio.ed.ac.uk/software/figtree/) (Table S4); genomes that did not have more than 1000 proteins matching with all the genomes were removed. Default parameters were used to run the phylogenetic reconstruction with 1000x bootstrap resampling analyses. The resistome of these genomes were curated from the Isolates Browser database of NCBI (https://www.ncbi.nlm.nih.gov/pathogens/isolates#/search/) (Tables S5-S6). The phylogeography of the resistant clones/clades per species was mapped manually unto an African map to show their geographical distribution.

## Results

### Characteristics of Included Studies

The literature search returned 1495 research articles: 1490 articles (from PubMed, Web of Science, ScienceDirect) and five articles from other sources obtained through manual search. Duplicates were removed and the remaining 868 non-duplicated articles’ titles and abstracts were screened. Full-text review of the remaining 309 manuscripts resulted in 146 studies being used for the qualitative and quantitative analyses (Figure 1) (Tables S1-S3).

The included articles spanned 31 countries from animal (A), human (H) and environmental (E) samples, with South Africa having the most (n=24) studies: **Algeria** (n=10; A=3, H=6, E=1); **Benin** (n=2; H=1, E=1); **Botswana** (n=2; A=1, H=1); **Burkina Faso** (n=7; A=1, H= 6,); **Cameroon** (n=3; A=1, H=2); **Central African Republic** (CAR)(n=2; H=2); **Chad** (n=1; H=1); **Cote D’Ivoire** (n=1; H=1); **Democratic Republic of Congo** (DRC) (n=2; H=1, E=1); **Egypt** (n=18; A=5, H=12, E=1); **Ethiopia** (n=5; A=2, H=3); **Gabon** (n=1; A=1); **Ghana** (n=5; A=1, H=4); **Kenya** (n=6; A=1, H=5); **Libya** (n=2; H=2); **Malawi** (n=1; H=1); **Mali** (n=1; H=1); **Morocco** (n=1; H=1); **Mozambique** (n=1; H=1); **Niger** (n=2; H=2); **Nigeria** (n=15; A=1, H=9, E=5); **Sao Tome & Principe** (n=1; H=1); **Senegal** (n=2; H=2); **Sierra Leone** (n=2; H=2); **South Africa** (n=35; A=4, H=24, E=7); **Tanzania** (n=10; A=3, H=6, E=1); **Tunisia** (n=10; A=3, H=7); **Uganda** (n=2; A=1, H=1); **Zambia** (n=2; A=1, H=1); **Zimbabwe** (n=2; A=1, H=1).

These studies involved 23,157 isolates from 65,119 specimens (isolation rate of 35.56%): 2,560 isolates from 5,950 animal specimens (43.03% isolation rate), 16,225 isolates from 57,464 human specimens (isolation rate of 28.24%), and 4,372 isolates from 1,705 environmental specimens (isolation rate of 256.42%). The various species identified per each study is summarised under the respective specimen source in Table 1; the per country breakdown is shown in Tables S1-S3.

**Table 1.** Species distribution frequencies per specimen sources

| Genera (species) | Animal specimens<br>(n = 5 950) | Human specimens<br>(n = 57 464) | Environmental<br>specimens (n = 1 705) | Total | Genomes included in<br>phylogenomics |
| --- | --- | --- | --- | --- | --- |
| *Escherichia (coli, hermannii, vulneris)* | 1566 | 4899 | 2827 | 9292 | 592 |
| *Salmonella (Enteritidis, Paratyphi, Typhi, Typhimurium)* | 500 | 1262 | 11 | 1773 | 487 |
| *Klebsiella (pneumoniae, oxytoca, varicola, michiganensis)* | 204 | 2528 | 44 | 2776 | 311 |
| *Pseudomonas (aeruginosa, fluorescens, otitidis, putida)* | 45 | 1366 | 87 | 1498 | 95 |
| *Acinetobacter (baumannii, calcoaceticus, haemolyticus)* | 2 | 611 | 92 | 705 | 21 |
| *Campylobacter (coli, jejuni)* | 210 | 175 | 105 | 490 | 13 |
| *Citrobacter (amalonaticus, freundii, koseri, farmer)* | 1 | 133 | 6 | 140 | 192 |
| *Enterobacter (aerogenes, amnigenus, cloacae, sakazakii)* | 21 | 459 | 0 | 480 | 60 |
| *Achromobacter xylosoxidans* | 1 | 0 | 0 | 1 | 0 |
| *Providencia (stuartii, rettgeri)* | 1 | 48 | 6 | 55 | 132 |
| *Proteus (mirabilis, vulgaris)* | 0 | 394 | 53 | 447 | 159 |
| *Serratia (marcescens, liquefasciens, odorifera)* | 0 | 144 | 4 | 148 | 197 |
| *Burkholderia spp.* | 0 | 6 | 0 | 6 | 7 |
| *Vibrio cholerae* | 0 | 24 | 279 | 303 | 180 |
| *Morganella (morganii)* | 0 | 31 | 19 | 50 | 85 |
| *Neisseria (meningitidis, gonorrhoea)* | 0 | 207 | 0 | 207 | 199 |
| *Bordetella (bronchiseptica)* | 0 | 1 | 2 | 3 | 21 |
| *Alcaligenes faecalis* | 0 | 3 | 47 | 50 | 0 |
| *Stenotrophomonas (maltophilia)* | 0 | 5 | 126 | 131 | 6 |
| *Leclercia adecarboxylata* | 0 | 1 | 0 | 1 | 0 |
| *Pantoea (dispersa, agglomerans)* | 0 | 2 | 1 | 3 | 21 |
| *Mycoplasma genitalium* | 0 | 266 | 0 | 266 | 23 |
| *Raoultella* | 0 | 3 | 0 | 3 | 0 |
| *Aeromonas* | 0 | 0 | 11 | 11 | 3 |
| *Bacillus* | 0 | 0 | 97 | 97 | 96 |
| *Brevundimonas* | 0 | 0 | 3 | 3 | 120 |
| *Chromobacterium* | 0 | 0 | 6 | 6 | 0 |
| *Myroides* | 0 | 0 | 2 | 2 | 0 |
| *Psychrobacter* | 0 | 0 | 3 | 3 | 0 |
| *Trabulsiella* | 0 | 0 | 1 | 1 | 8 |
| **Total** | **2551** | **12568** | **3832** | **<u>18951</u>** | **3028** |

- Discrepancies between total isolates in Table 1 and total isolates under Results arise from the fact that not all isolates were Gram-negative isolates. Non-Gram-negative isolates are not included in Table 1.

The 3028 genomes included in this study were also obtained from animals, humans, plants and the environment from 41 African countries: Angola, Benin, Botswana, Burkina Faso, Cameroon, CAR, Chad, Comoros, DRC, Djibouti, Egypt, Eritrea, Ethiopia, Gambia, Ghana, Guinea, Guinea-Bissau, Kenya, Lesotho, Madagascar, Malawi, Mali, Mauritania, Mauritius, Morocco, Mozambique, Namibia, Niger, Nigeria, Republic of the Congo, Rwanda, Senegal, Sierra Leone, South Africa, Sudan, Tanzania, Togo, Tunisia, Uganda, Zambia, and Zimbabwe (Table S6; Figures 2-8; Figures S7-S25).

**Figure 2.**
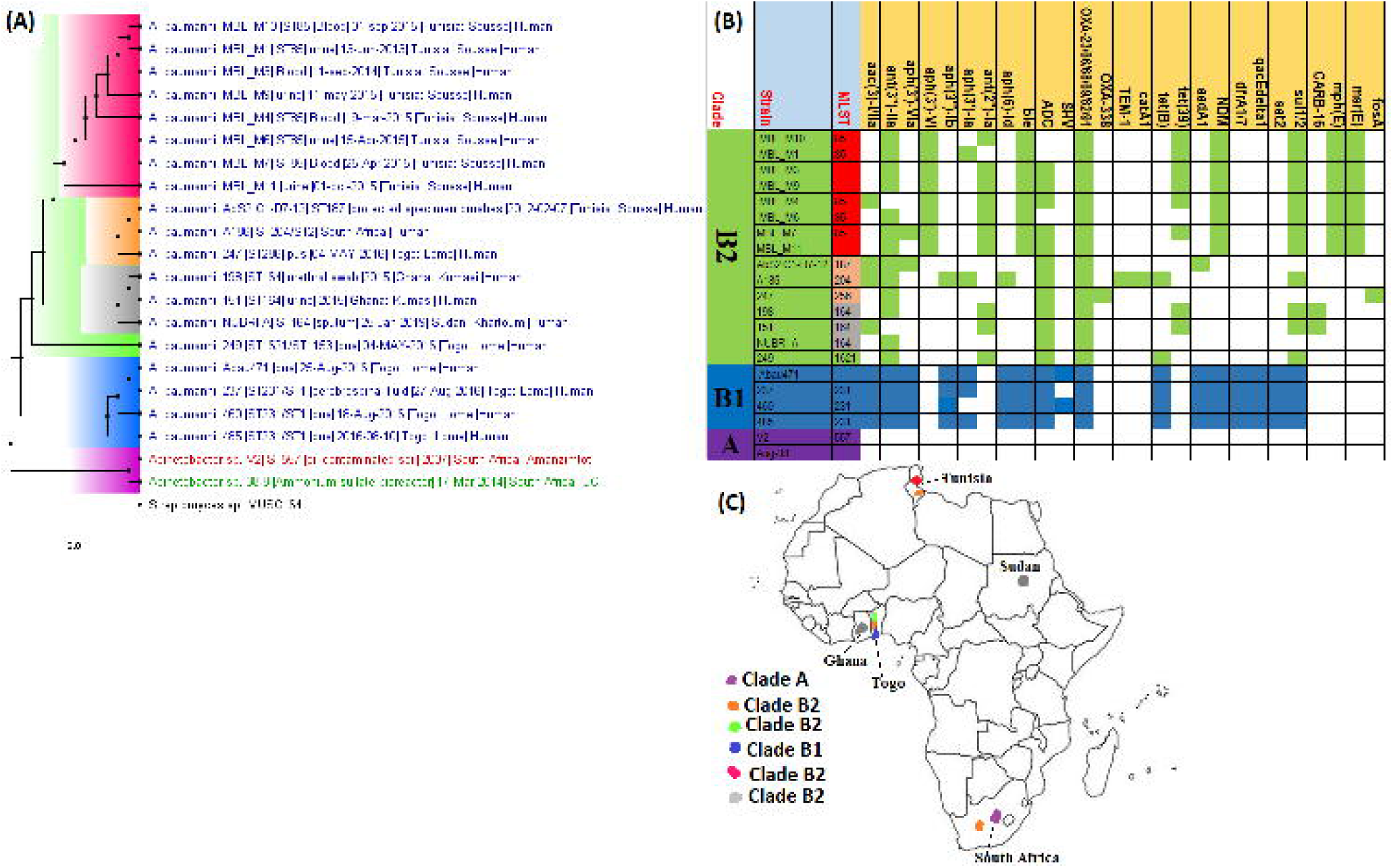
Phylogeographic distribution of *Acinetobacter baumannii* clades and associated resistomes in Africa. The included *A. baumannii* genomes were mainly from Tunisia, Togo, Ghana, Sudan, and South Africa, with clade-specific ARGs; most of these strains were from humans. Cluster A, which was not *A. baumannii*, had no ARGs, whilst clusters/clades B1 and B2 had OXA-23-/66-like carbapenemases, *ble, ant(2’’)-Id, aph(3’)-Ib* and *ant(3”)-IIa*. Isolates from humans, animals, the environment and plants are respectively coloured as blue, red, mauve/pink and green on the phylogeny tree.

### Species distribution (from included articles)

Of the 30 species isolated from the various human, animal and environmental specimens included in the studies used for this meta-analysis, the commonest were *Escherichia spp*. (n=9292), *Klebsiella spp*. (n=2776), *Salmonella enterica* (n=1773), *Pseudomonas spp*. (n=1498), and *Acinetobacter spp*. (n=705), which were all commonly isolated from human samples than from animal or environmental specimens; these statistics were also largely reflected in the species distribution in the genomics data (Table 1). These common pathogens, including *Neisseria gonorrhoea/meningitidis, Proteus mirabilis* and *Enterobacter spp*., were mostly concentrated in Algeria, Burkina Faso, Egypt, Ghana, Kenya, Libya, South Africa, Tanzania and Tunisia in humans (Fig. S1). South Africa, Tanzania and Nigeria, reported the highest concentrations of environmental species. Notably, *E. coli* and *S. enterica*, and to a lesser extent *Campylobacter coli/jejuni, Klebsiella spp*., and *Pseudomonas spp*., were the most isolated species from animals in the reporting countries. It is interesting to note that *N. gonorrhoea/meningitidis* were mainly reported from humans in Kenya and Niger whilst *Vibrio cholerae/spp*. were mostly isolated from the environment in South Africa and to a lesser extent, from humans in Cameroon (Fig. S1); yet, genomes of *N. meningitidis* were obtained from 10 countries in Southern, Eastern, Western and Northern Africa (Fig. 8).

*E. coli* was isolated in very high frequencies in almost all reporting countries except Kenya, Ethiopia, Botswana, Zambia and Senegal (in humans), Ghana, Burkina Faso and Botswana (in Animals) and Nigeria, Egypt and Cameroon (in the environment). *K. pneumoniae* was less common in humans in Egypt, Ethiopia, DRC, Cameroon, Botswana, Benin, Zimbabwe, Zambia, Niger and Malawi; it was hardly reported from animals in Egypt and Cameroon and only found in the environment in Nigeria. *S. enterica* was mainly distributed in Algeria, Ethiopia, Ghana, Kenya, and Zambia (in humans), and Zambia, Tunisia, S. Africa, Kenya, Ethiopia and Algeria (in animals); it was only reported from Egypt from the environment. In humans, *P. aeruginosa* was mostly found in Egypt, B. Faso, Tanzania, S. Africa and Nigeria whilst Egypt alone reported it in animals and Nigeria alone reported *Pseudomonas spp*. in the environment. Interestingly, *A. baumannii* was mainly concentrated in Ethiopia and Egypt (humans), and in Algeria (animals); *A. calcoaceticus/spp*. was found from the environment in South Africa and Nigeria (Fig. S1-S2).

Only *Campylobacter coli/jejuni* had more animal sources (n=210) than human (n=175) and environmental (n=105) sources and *S. enterica* was the second most isolated species from animal specimens after *E. coli* (Fig. S2). Notably, *Vibrio cholerae* (n=279), *Stenotrophomonas maltophilia* (n=126), *Bacillus spp*. (n=97), *Alcaligenes faecalis* (n=47), *Aeromonas spp*. (n=11), *Chromobacterium spp*. (n=6), *Brevundimonas spp*. (n=3), *Psychrobacter spp*. (n=3), *Myroides spp*. (n=2), and *Trabusiella spp* (n=1) were either mainly or only found from environmental sources (Table 1).

*Neisseria meningitidis/gonorrhoea* and *Mycoplasma genitalium*, two sexually transmitted infectious pathogenic species, were mainly found in clinical samples (Table 1; Fig 5). However, other *Mycoplasma spp*. were found in chicken (*M. gallinarum/gallinaceum/pullorum*), goat (*M. bovis*), cattle (*M. mycoides*), ostrich (*M. nasistruthionis*), sewage (*M. arginini*) and humans (*M. pneumoniae*), and these had no known resistance genes (Fig. S19). Species such as *Burkholderia cepacia/spp*., *Bordetella spp*., *Morganella morganii, A. faecalis, S. maltophilia, Leclercia adecarboxylata, Pantoea spp*. and *Raoultella spp*. were rare in clinical samples (Table 1; Fig. S14, S20 & S24).

### Geographical and host distribution of clones, ARGs and MGEs

The clonality of *A. baumannii, C. coli/jejuni, E. coli, K. pneumoniae, P. aeruginosa, V. cholerae, S. marcescens, N. gonorrhoeae/meningitidis*, and *S*. Entiritidis/Typhi/Typhimurium strains were reported from the various countries out of the 30 species (Fig. S3). However, only *E. coli* clones viz., ST38, ST69, ST131, ST410 etc. and groups A/B/C/D were found in humans (in Algeria, B. Faso, CAR, Egypt, Libya, Nigeria, Sao Tome & Principe, Tanzania, Tunisia and Zimbabwe), animals (Algeria, Egypt, Ghana, Tunisia and Uganda) and the environment (Algeria and South Africa). Specifically, *E. coli* ST38 was found in humans (Algeria) and animals (Ghana) and groups A/B/D were found in humans (Egypt), animals (Algeria, Egypt, Tunisia, Uganda and Zimbabwe) and the environment (Algeria and South Africa) (Fig. S3). Inter-country detection of *E. coli* ST131 in humans was also observed in Algeria, B. Faso, CAR, DRC, Tanzania, Tunisia and Zimbabwe. *K. pneumoniae* ST101 was also found in Algeria, South Africa, and Tunisia. As well, multiclonal *C. jejuni* strains (i.e. ST19, ST440, ST638, ST9024 etc.) were found in humans and animals from Botswana (Fig. S3).

The clones of the various species from the genomic data did not always agree with that obtained from the included articles in terms of geographical distribution and incidence (Fig. S3 & Table S6). For instance, the *E. coli* genomes were highly multiclonal, consisting of 202 clones; the commonest of these were ST661 (n=56), ST10 (n=42), ST443 (n=35), ST131 (n=24), and ST29 (n=10). *K. pneumoniae* (85 clones) and *S. enterica* (66 clones) genomes were also very multiclonal, with *K. pneumoniae* ST101 (n=40), ST152 (n=22), ST15 (n=16), ST14 (n=12), ST17 (n=12), ST147 (n=10) and *S. enterica* ST2 (n=148), ST1 (n=137), ST198 (n=20), ST11 (n=12), ST313 (n=12), ST321 (n=10) and ST2235 (n=10) being very common. Notably, *N. meningitidis* (genomes) ST11 (n=45), ST2859 (n=22), ST1 (n=9), ST2859 (n=9) etc. were also common in humans from Ghana (n=63 isolates), B. Faso (n=57 isolates), Niger (n=28 isolates) etc. as seen in the articles (Fig. S3 & Table S6). Contrarily, *P. aeruginosa* (ST234 & ST235), *A. baumannii* (ST1, ST85 & ST164), *C. jejuni* (ST362), *V. cholerae* (ST69 & ST515), *Bordetalla pertussis* (ST1 & ST2 in Kenya), *Mycoplasma pneumoniae* (in Egypt and Kenya), *Bacillus cereus* and *Bacillus subtilis* (genomes) had relatively few dominant clones (Table S6).

ARGs mediating resistance to almost all known Gram-negative bacterial antibiotics were found in the included articles, with most of these ARGs being isolated from human strains than animal and environmental species in a descending order (Fig. S4). Notably, ARGs conferring resistance to β-lactams, specifically ESBLs such as CTX-M, TEM, SHV, OXA, GES and AmpCs such as CMY, FOX, DHA, FOX, MOX, ACC, EBC and LEN were commonly identified in human, animal and environmental isolates from most countries, with *bla*_CTX-M_ and *bla*_TEM_ being the most identified ARGs (Fig. S2 & S4). Notably, OXA and GES ESBLs and all the AmpCs as well as carbapenemases (i.e., OXA-48/181/204, OXA-23/51/53, NDM, IMP, SPM, VIM, KPC, GES-5) were not reported from animal or environmental isolates; only OXA-61 (from *C. jejuni*) was found in animal isolates in Botswana. Carbapenemase genes were relatively less detected in human strains and reported from a few countries: the metallo-β-lactamases such as NDM, IMP, SPM and VIM were mainly found in Egypt, South Africa, Tanzania, Tunisia, and Uganda; KPC and GES-5 were common in South Africa and Uganda; and the OXA-types were found in Algeria, Egypt, Nigeria, Sao Tome & Principe, South Africa, Tunisia and Uganda (Fig. S4).

Second to the β-lactams, there were frequent detection of diverse fluoroquinolone resistance mechanisms in human and animal isolates from almost all the countries: *aac(6’)-Ib-cr, aac(3’)-IIa, aac(3’)-Ih, QnrA/B/D/S, OqxAB*, chromosomal mutations in *gyrAB* and *parCE* and *qepA* in a descending order of frequency. None of these mechanisms were found in environmental strains. Moreover, aminoglycosides resistance mechanisms, including *aac(6’)-Ib-cr* that also confers resistance to fluoroquinolones, were equally highly distributed in human and animal isolates, with relatively limited occurrence in environmental strains. Among these aminoglycoside mechanisms were *aadA, strAB, aph(3’), aph(6’), ant(2’), ant(3’)*, and the 16S rRNA methyltransferases such as *rmtC/F* and *armA*.

Other common resistance mechanisms that were highly distributed in almost all strains from almost all the included countries were *sul1/2/3* and *dfrA* (mediating resistance to sulpha-methoxazole-trimethoprim); these were mostly found in animal and human strains and relatively less isolated in environmental strains. Chloramphenicol resistance genes viz., *cmlA/B* and *catA/B*, were also found in animal and human isolates in substantial numbers whilst ARGs for florfenicol (*floR*) and fosfomycin (*fosA*) were very rare, being found in only human strains. Of note, tetracycline ARGs, *tet*(O/A/B/C/D/G/K/M39), were almost fairly distributed in strains from humans, animals and the environment in relatively few countries such as Algeria (E), Botswana (A & H), Cameroon (E), Ethiopia (A), Malawi (H), Nigeria (E), South Africa (A, H, E), Tanzania (A, E), Tunisia (A, H), Uganda (A), and Zambia (A).

Interestingly, colistin resistance mechanisms such as *mcr-1* and chromosomal mutations in *pmrAB* were very rare. Particularly, *pmrAB* mutations were only recorded in human strains from Tunisia whilst *mcr-1* genes were only reported in South Africa (A, H, E), Sao Tome & Principe (H), and Tunisia (A). Other rare ARGs, found mainly in human isolates, included *blaZ, pse-1*, and *penA* (conferring penicillin resistance), *ermABC* and *mph(A)* (encodes erythromycin/macrolides resistance), *cmeAB* (multidrug efflux system in *Campylobacter jejuni*), *porAB* (porin in *Neisseria spp*.), *macA/B* (encodes part of the tripartite efflux system MacAB-TolC for transporting macrolides from the cytosol), *qacE*Δ*1* (encodes resistance to quarternary ammonium compounds through efflux), and *mexAB* (encodes multidrug resistance (MDR) efflux pumps in *Pseudomonas spp*.).

These ARGs were found associated with MGEs such as plasmids, integrons, insertion sequences (ISs) and transposons, mainly in human isolates; MGEs in animal and environmental isolates were rarely described (Fig. S5-S6). The commonest MGEs were IncF-type, IncX_3_, ColRNAI, IncR, IncY, IncL/M, A/C, IncH, and IncQ plasmids, *IS*EcP1 and class 1 integrons (IntI1). ColE, IncU, IncLVPK, IncP, IncI, IncN, ColpVc, and ColKp3 plasmids, *Tn*2006, *IS*Kpn19, *Tn*3, *IS*26, *Tn*21 and *IS*AbaI transposons and ISs were less identified MGEs. Specifically, IncF-type plasmids were commonly associated with ESBLs and carbapenemase genes whilst IntI1 was common with both β-lactamases and non-β-lactamase genes such as *sul-1/2/3, dfrA, catA/B, floR, qepA* and *qnrA/B/D/S. IS*EcP1 was common around *bla*_CTX-M-15_, *Tn*206 bracketed *bla*_OXA-23/51_ and *aac(3’)-I*, and *Tn*402-lik transposon harboured *bla*VIM-5, *aph(3’)-Ib, aph(6’)-Id, tet*(C/G), and *floR* (in environmental strains in Nigeria) (Fig. S5-S6).

### Resistance rates

Resistance rates to the various antibiotics were highest among human strains, particularly in Egypt, Ethiopia, Mali, Senegal, Tunisia and Uganda among Enterobacteriaceae such as *E. coli, K. pneumoniae, Salmonella enterica*, and *Providencia rettgeri/stuartii, Neisseria meningitidis/gonorrhoea, P. aeruginosa*, and *A. baumannii*. These human strains expressed very high resistance to almost all the antibiotic classes including the aminoglycosides, β-lactams, fluoroquinolones, tetracyclines, sulphamethoxazole-trimethoprim (SXT) and phenicols. Comparatively, strains from animals and the environment expressed lesser resistance rates. Specifically, *A. baumannii, E. coli, Salmonella enterica*, and *Providencia spp*., were resistant to ampicillin, amikacin, chloramphenicol, kanamycin, tetracycline, streptomycin, sulfonamide and SXT in most of the included countries. Notably, the resistance rates of environmental *E. coli, S. enterica, K. pneumoniae/oxytoca*, and *Citrobacter freundii/koseri* strains to fluoroquinolones, tetracycline, sulphonamide, SXT, and ceftriaxone were substantially high in Algeria, Benin and Egypt (Tables S1-S3).

### Phylogenomic and resistome analyses: evolutionary epidemiology of resistance

Phylogenetically, strains belonging to the same clades were found in different countries, and in a limited measure, in humans, animals, the environment and in plants. Among the species, certain countries contained only a single clade of a species whilst some countries contained several clades of the same species: *E coli* (Algeria), *K. pneumoniae* (Mali), N. meningitis (South Africa, DRC), *Campylobacter spp*. (South Africa) etc (Fig. 3-8, S7-S25). Within specific clades were found, in a few cases, isolates from different sample sources: clades A, B and C (*E. coli*), clade B (*P. aeruginosa*) etc. (Fig. 3 & 6, S7). Notably, strains belonging to different MLSTs were found within the same clades. Generally, the genomes of included species were from Southern, Eastern, Western and Northern Africa, with little or none from Central Africa; countries reporting most genomes included Ghana, Mali, Nigeria, Cameroon, Tunisia, Algeria, Egypt, Kenya, Tanzania, Mozambique and South Africa (Fig. 3-8, S7-S25).

**Figure 3.**
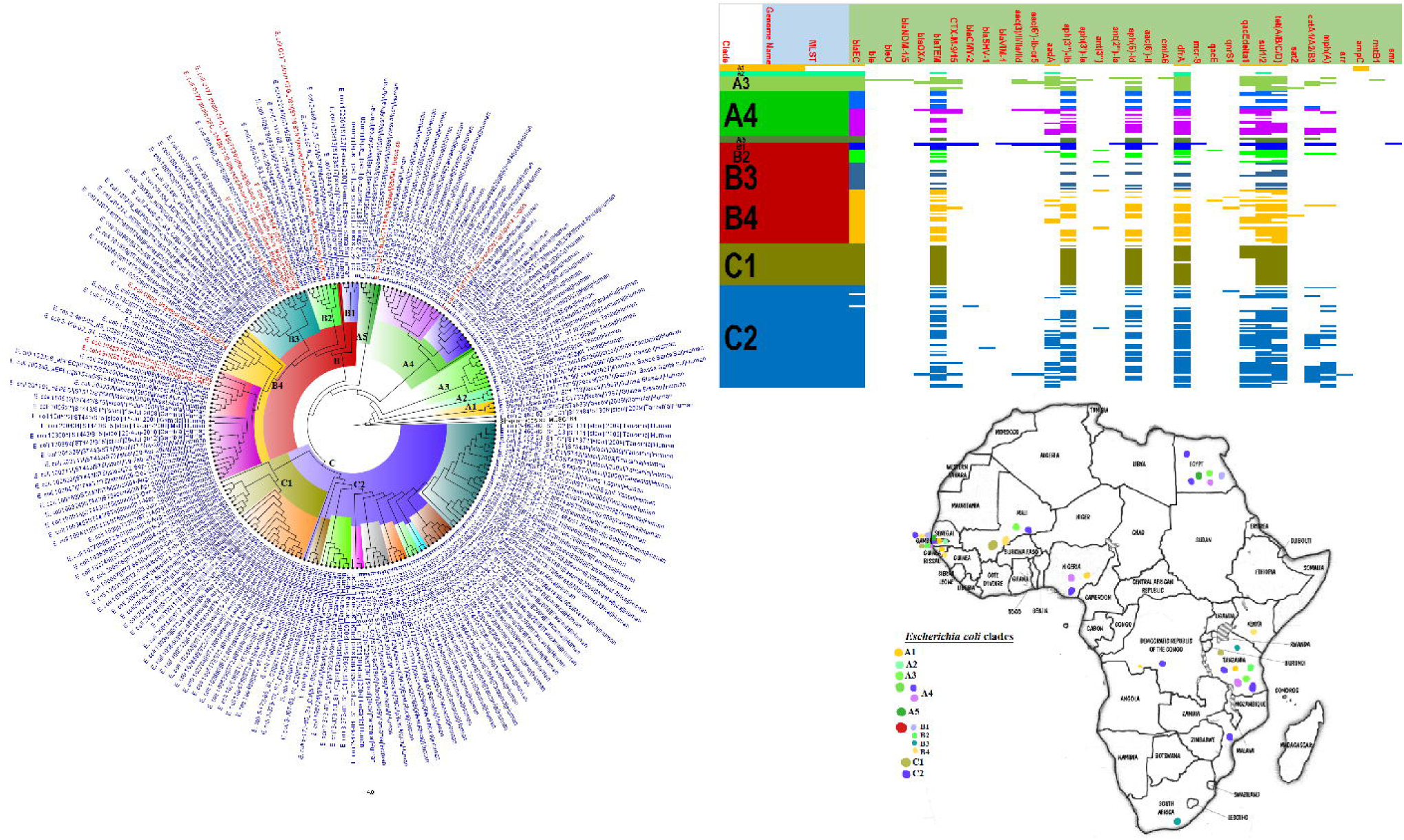
Phylogeographic distribution of *Escherichia coli* clades and associated resistomes in Africa. The *E. coli* clades were mostly from humans, mainly distributed in West and East Africa, Egypt and South Africa. Relatively few were from the environment and animals. The ARGs (*tet (A/B/C/D), bla*_TEM-1_, *sul, aph(3”)-Ib, aph(6)-Id*, and *dfrA*) were mostly conserved across the various clades, which were not region-specific, but mixed up. Strains from humans shared very close phyletic relationship with strains from animals and plants. Inter-country as well as human-animal dissemination of isolates of the same clade were observed. Isolates from humans, animals, the environment and plants are respectively coloured as blue, red, mauve/pink and green on the phylogeny tree.

*Enterobacter spp*. and *K pneumoniae* strains contained the largest and richest repertoire of resistomes (Fig. 4, S8 & S17). In a descending order, *S. marcescens, S. enterica, E. coli, A. baumannii, P. aeruginosa, V. cholerae, Citrobacter freundii, Providentia rettgerii, Proteus mirabilis* and *M. morganii* strains harboured a rich and diverse repertoire of ARGs. No known ARGs were found in the other species, although the literature reported resistance mechanisms in *N. meningitidis* (*gyrA, penA*, and *rpoB*), *N. gonorrhoeae* (*gyrA, penA, ponA, mtrR, porB* and *bla*_TEM_), *C. jejuni/coli* (*bla*_OXA-61_, *tet*(O), *gyrA*(T**86**I), *cmeABC*, and *aph(3)-I*), *M. genitalium* (*gyrA* and *parC*) and *S. maltophilia* (*sul3*) (Tables S1-S3). Some ARGs were conserved within specific clades and species: NDM-1 (*S. marcescens*), OXA-23/66-like (*A. baumannii*), TEM-1 (*E. coli, K. pneumoniae*), CTX-M (*Enterobacter spp*. and *K. pneumoniae*), SUL-1/2/3 (*Enterobacter spp*., *E. coli, K. pneumoniae, S. enterica*), dfrA (*Enterobacter spp*., *E. coli, K. pneumoniae, S. enterica*), QnrB/S (*Enterobacter spp*., *K. pneumoniae*), TET (*Enterobacter spp*., *E. coli, N. meningitidis, K. pneumoniae, S. enterica*) etc. (Fig. 3-8, S7-S25).

**Figure 4.**
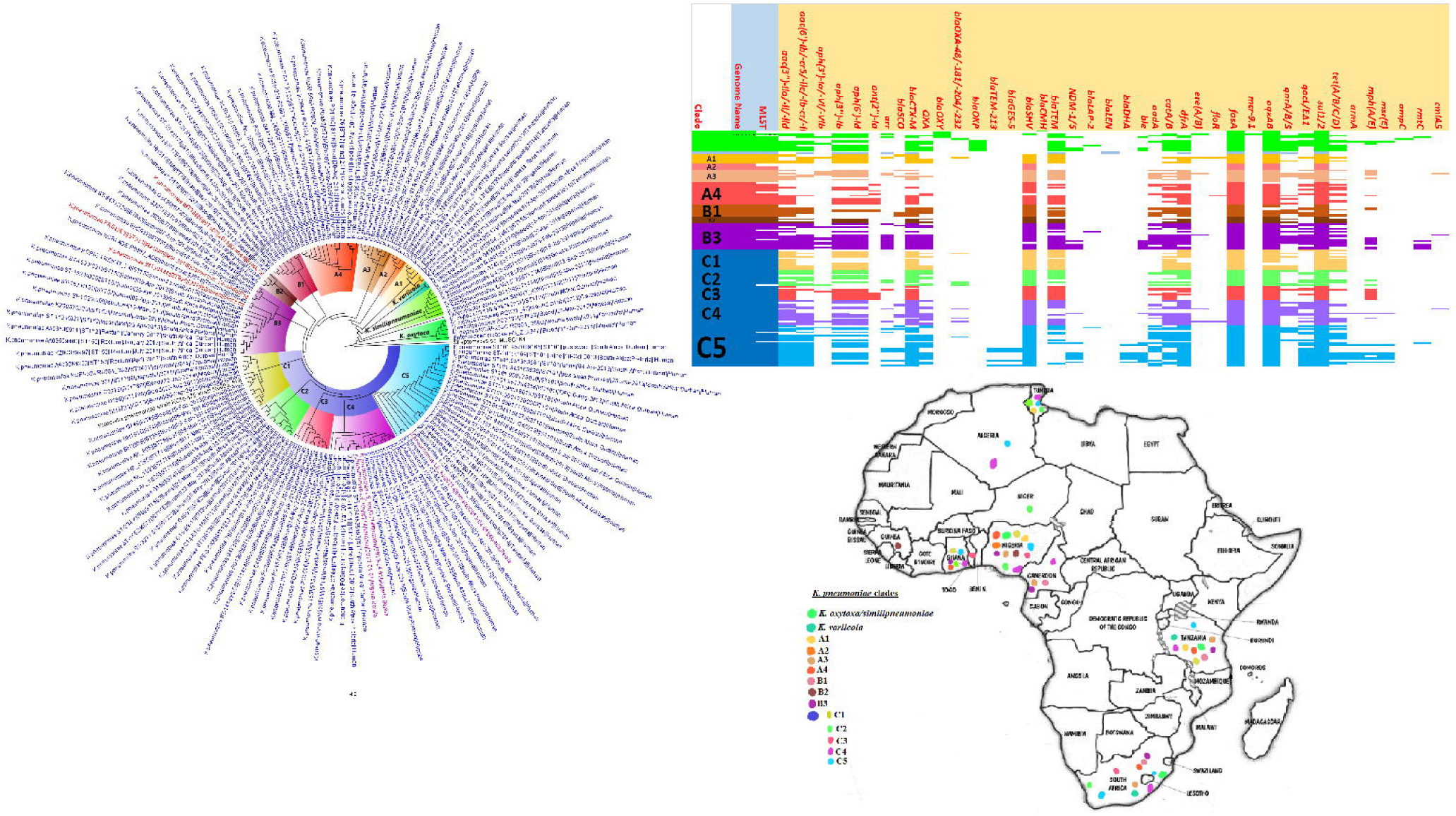
Phylogeographic distribution of *Klebsiella pneumoniae* clades and associated resistomes in Africa. *Klebsiella pneumoniae* was mainly from humans, with a few being isolated from plants and animals. The various clades were mixed up in South Africa, Tanzania, Uganda, West and North Africa. Strains from humans shared very close phyletic relationship with strains from animals and plants. The clades harboured many conserved ARGs (n=14): *aac(3’)-Ia/IIa, aac(6’)-IIa/Ib-cr, aph(3”)-Ib, aph(6’)-Id, bla*_CTX-M_, *bla*_OXA_, *bla*_SHV_, *bla*_TEM_, *aadA, catA/B, dfrA, fosA, oqxAB, sul1/2*. Other ARGs that were substantially found in *K. pneumoniae* included *arr, mph(A/E), qnrA/B/S, qacE*Δ*1* and *bla*_NDM-1/5._ A few genes were restricted to certain clades. Inter-country as well as human-animal dissemination of isolates of the same clade were observed. Isolates from humans, animals, the environment and plants are respectively coloured as blue, red, mauve/pink and green on the phylogeny tree.

### Diagnostics

Phenotypic and molecular methods were employed by the included studies to determine the species identity, antibiotic sensitivity (AST), genotype/clone and resistance mechanisms of the isolates. Broth microdilution (BMD) and disc diffusion methods were common phenotypic tests used for determining the AST of the isolates. Vitek, E-test and agar dilution methods were less used. PCR or PCR-based typing methods such as multi-locus sequence typing (MLST), repetitive element PCR (REP), and enterobacterial repetitive intergenic consensus (ERIC)-PCR were more commonly used to determine the clonality of Enterobacterial isolates than non-PCR-based techniques such as pulse-field gel electrophoresis (PFGE), which is more laborious. Finally, PCR was the commonest tool used for determining the ARGs of the isolates, with the use of whole-genome sequencing being limited (Table 2).

**Table 2.** Diagnostics used for detection, typing and resistance characterisation

| Diagnostic or typing tool/assay | Animals | Humans | Environment |
| --- | --- | --- | --- |
| Phenotypic antibiotic sensitivity testing |  |  |  |
| Agar dilution method | 1 | 0 | 3 |
| Disk diffusion method | 19 | 35 | 7 |
| Broth Micro-dilution MIC test | 12 | 51 | 10 |
| E-test | 0 | 7 | 1 |
| VITEK®2 System | 0 | 19 |  |
| Molecular typing |  |  |  |
| AFLP<sup>1</sup> | 1 | 0 | 0 |
| MLST/cgMLST<sup>2</sup> | 7 | 47 | 2 |
| Whole-genome sequencing (WGS) | 3 | 6 | 0 |
| PCR-based phylogenetic typing (ERIC-/ | 9 | 11 | 3 |

<sup>1</sup> Amplified fragment length polymorphism (AFLP)
<sup>2</sup> Multi-locus sequence typing (MLST)/core-genome MLST (cgMLST)

|  |  |  |  |
| --- | --- | --- | --- |
| REP-PCR) |  |  |  |
| PFGE/S1-PFGE<sup>3</sup> | 3 | 18 | 1 |
| Molecular detection of ARGs (antibiotic resistance genes) |  |  |  |
| PCR | 27 | 92 | 19 |
| WGS | 5 | 23 | 1 |
| Sanger sequencing | 0 | 0 | 1 |
| Southern blotting | 0 | 2 | 0 |

## Discussion

The resistome dynamics or epidemiology, phylogenomics and phylogeography of Gram-negative bacterial species and associated clones, MGEs and ARGS in Africa are herein limned for the first time. We show that *E. coli, K. pneumoniae, S. enterica, A. baumannii, P. aeruginosa, N. meningitidis/gonorrhoeae, V. cholerae, S. marcescens, Enterobacter spp. C. jejuni, Mycoplasma spp*., *Providencia spp*., *Proteus mirabilis and Citrobacter spp*. are major pathogenic species with rich and diverse resistomes and mobilomes, circulating in humans, animals and environment in Southern, Eastern, Western and Northern Africa. The poorer sanitary conditions, food insecurity as well as weaker healthcare and diagnostic laboratory capacities in Africa are well-known, accounting for the higher infectious disease rates on the continent ^2–5,18,24^. Subsequently, the combination of diverse ARGs in highly pathogenic species with wide geographical distribution on the continent is a cause for concern as it provides fertile breeding grounds for periodic and large outbreaks with untold morbidities and mortalities ^25–28^.

Although *E. coli* was the most isolated species in human, animal and environmental specimens, it did not harbour the most diverse and richest resistome repertoire. This is interesting as *E. coli* is mostly used as a model and index organism to study Gram-negative bacterial resistance epidemiology ^18,29^. Whereas the higher numbers of *E. coli* strains obtained in Africa could be due to its easy cultivability, identification, and exchange of resistance determinants ^17,30^, as well as its common use as a model and/or index organism^18,29^, its lower resistome diversity and richness could mean it is not representative of the actual resistome circulating in any niche at a given point in time. Hence, *Enterobacter cloacae/spp*. and *K. pneumoniae*, which contained richer resistomes, could serve as better representatives and reporters of prevailing ARGs in any niche at a point in time. Particularly, the richer resistomes of these two species strongly suggest that they can easily exchange ARGs between themselves and other species, as reported already ^17,30–32^.

The higher resistome diversity, as well as the high isolation rates, of *K. pneumoniae* and *Enterobacter spp*. are not surprising. Specifically, *K. pneumoniae* is the most isolated clinical bacterial pathogen in many countries worldwide, found to be involved in many fatal and multidrug resistant infections ^25,28,33,34^. International clones such as *K. pneumoniae* ST208 and ST101 are implicated in the clonal dissemination of carbapenemases as well as colistin and multidrug resistance ^25,28,33–36^; although ST208 was absent in Africa, ST101 was common in several countries. As well, *Enterobacter spp*. are increasingly being isolated in many clinical infections in which they are found to as a major host of *mcr* colistin resistance genes and other clinically important MDR determinants ^34,37–39^. Notwithstanding its lower resistome diversity compared to *Enterobacter spp*. and *K. pneumoniae*, the *E. coli* isolates contained important ARGs, such as *mcr-1, bla*_NDM-1_, *bla*_OXA-48/181_, and *bla*_CTX-M-15_ (Tables S1-S3), which can be transferred to other intestinal pathogens ^16,17,30^. Notably, the *E. coli* isolates also exhibited high resistance rates against important clinical antibiotics (Tables S1-S3). Finally, the *Enterobacter spp*., *K. pneumoniae*, and *E. coli* strains were generally highly multi-clonal and evolutionarily distant, suggesting little clonal dissemination (except *E. coli* ST103 and *K. pneumoniae* ST101) of these species across the continent, albeit local and limited inter-country outbreaks were observed (Fig. 2-3, S7-S8 & S17) ^33,40^.

*S. enterica* and *C. coli/jejuni*, which are important zoonotic and food-borne pathogens ^24,41^, were found in animal/food, human and environmental specimens, although *S. enterica* was more common and had a higher resistome diversity than *C. coli/jejuni* (Tables 1 & S1-S3; Fig. 7, S9 & S15). Notably, *S*. Typhi, *S*. Typhimurium, *S*. Entiritidis and *S*. Bovismorbificans were mostly isolated from humans, with some *S*. Entiritidis and *S*. Infantis strains being isolated from both humans and cattle (Fig. 7 & S9). Indeed, reports of *S*. Typhimurium and *S*. Entiritidis isolation from pigs and poultry respectively, as well as their implication in fatal zoonotic infections through contaminated food animal consumption, are well-documented^24,42–45^. *S*. Typhi, a common food-borne pathogen that infects millions of people worldwide annually and results in typhoid fever, diarrhoea and death in severe cases ^18,20,21,46^, was the third most common species to be isolated and the fourth species to host the most resistome repertoire. As shown in Tables S4-S6 and Fig. 7 & S9, several isolates from Eastern, Southern and Western Africa shared the same clone (ST1, ST2) and clade, representing clonal outbreaks affecting many people over a large swathe of Africa. Moreover, *S*. Typhi and other *S. enterica* serovars recorded very high resistance rates to first-line antibiotics in many African countries within the study period (Table S2). This is a very concerning observation given the widespread and periodic incidence of outbreaks involving this pathogen in most developing countries ^18,21^.

**Figure 5.**
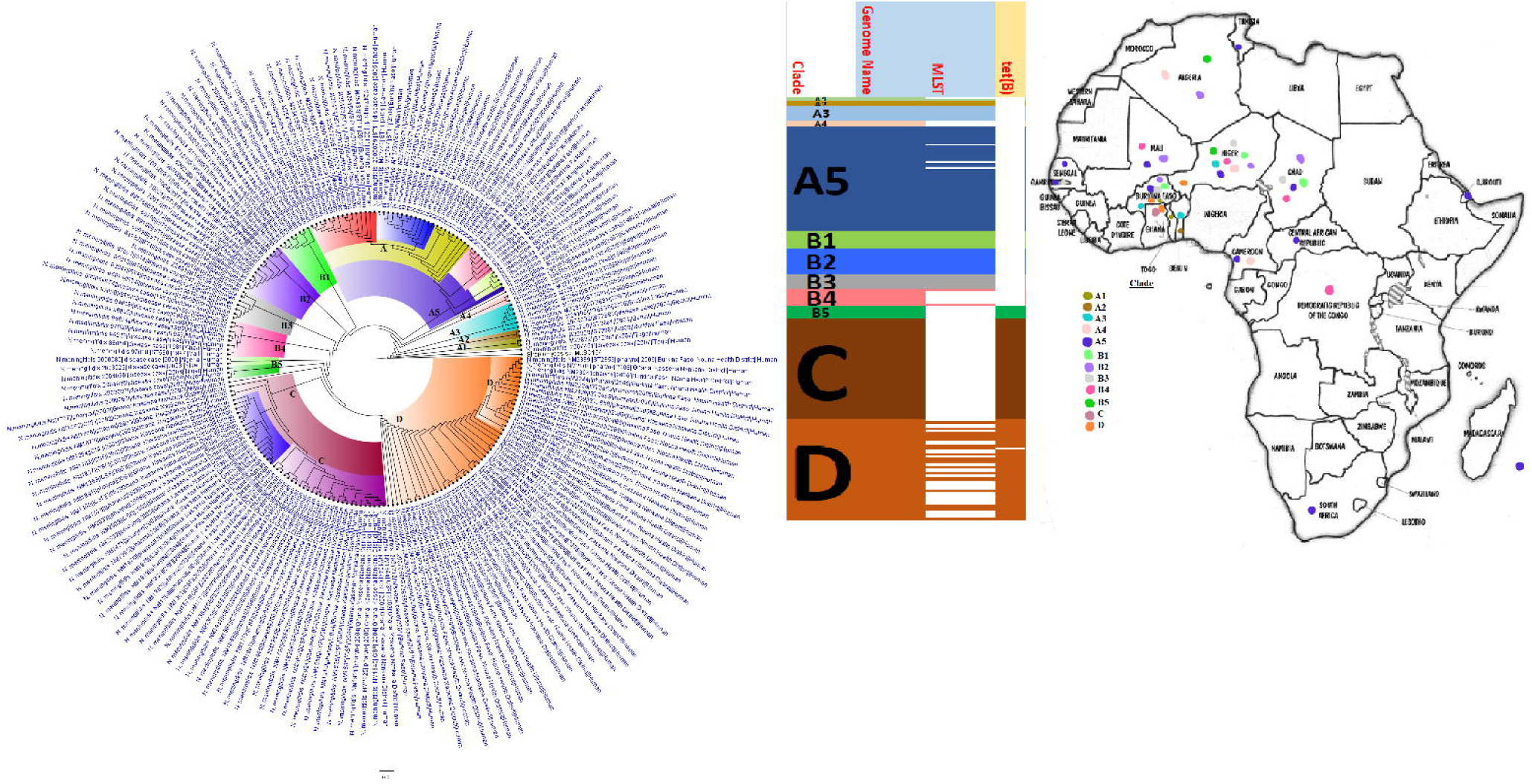
Phylogeographic distribution of *Neisseria meningitidis* clades and associated resistomes in Africa. *Neisseria meningitidis* was only from human samples from South Africa, Comoros, DRC, CAR, Cameroon, West Africa, Djibouti and Algeria; however, *N. meningitidis* was most concentrated in West Africa and the ARG (only *tet(*B)) was only found in clades B5, C and D. Inter-country dissemination of isolates of the same clade were observed. Isolates from humans, animals, the environment and plants are respectively coloured as blue, red, mauve/pink and green on the phylogeny tree.

**Figure 6.**
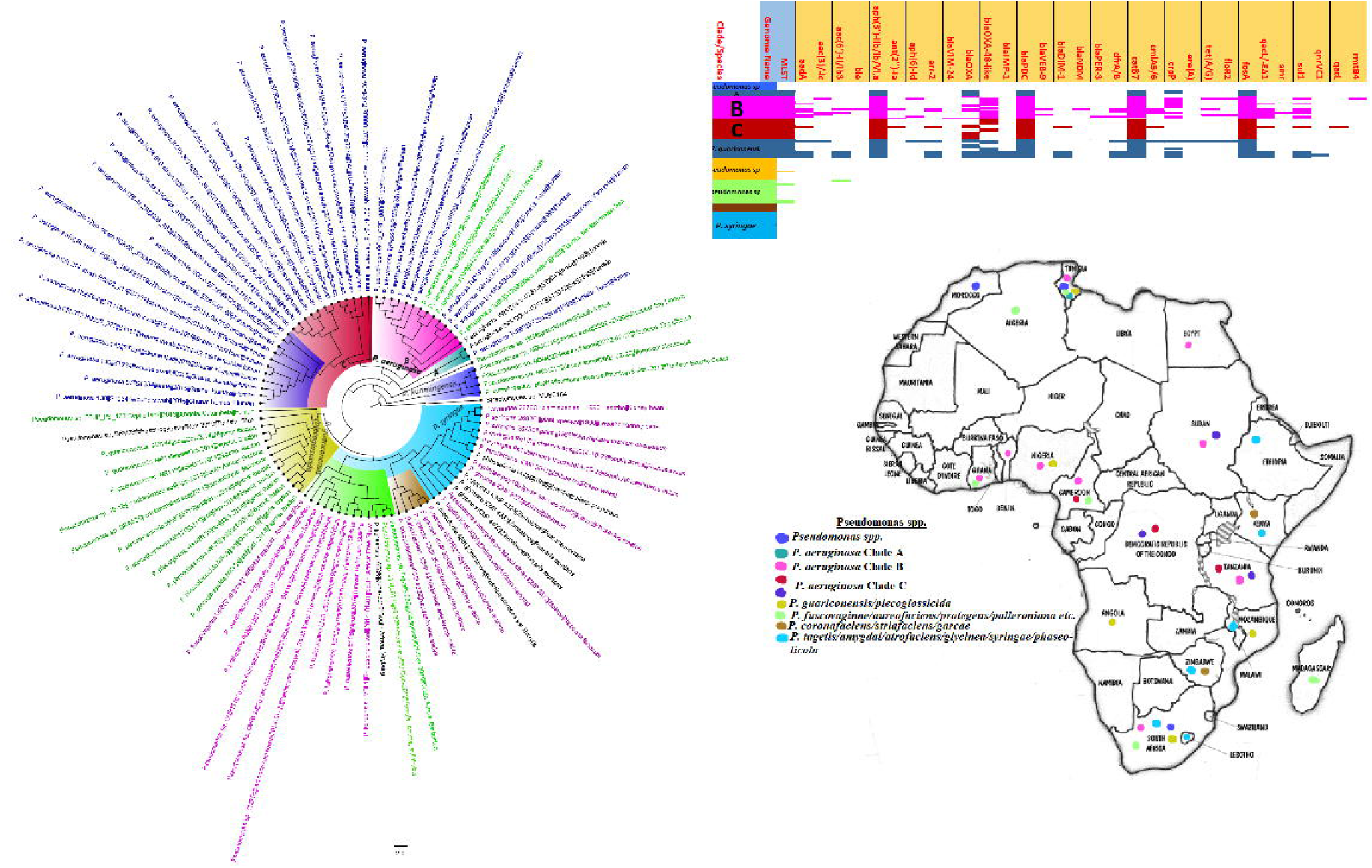
Phylogeographic distribution of *Pseudomonas spp*. clades and associated resistomes in Africa. *Pseudomonas spp*. were widely distributed in Africa, particularly Southern and Eastern Africa, DRC, West and (Cameroon, Nigeria, Benin, Ghana) North (Morocco, Algeria, Tunisia and Egypt) Africa. The other *Pseudomonas spp*. were mainly found in the environment and plants with no ARGs whilst *P. aeruginosa* were found in humans and the environment with *aph(3’)-IIb, bla*_OXA_, *bla*_PDC_, *catB7* and *fosA* being largely conserved in this pathogen. Clade B had additionally conserved genes such as *sul1/2*, and *qacL/*Δ*E1*. Inter-country as well as human-environment dissemination of isolates of the same clade were observed.Isolates from humans, animals, the environment and plants are respectively coloured as blue, red, mauve/pink and green on the phylogeny tree.

**Figure 7.**
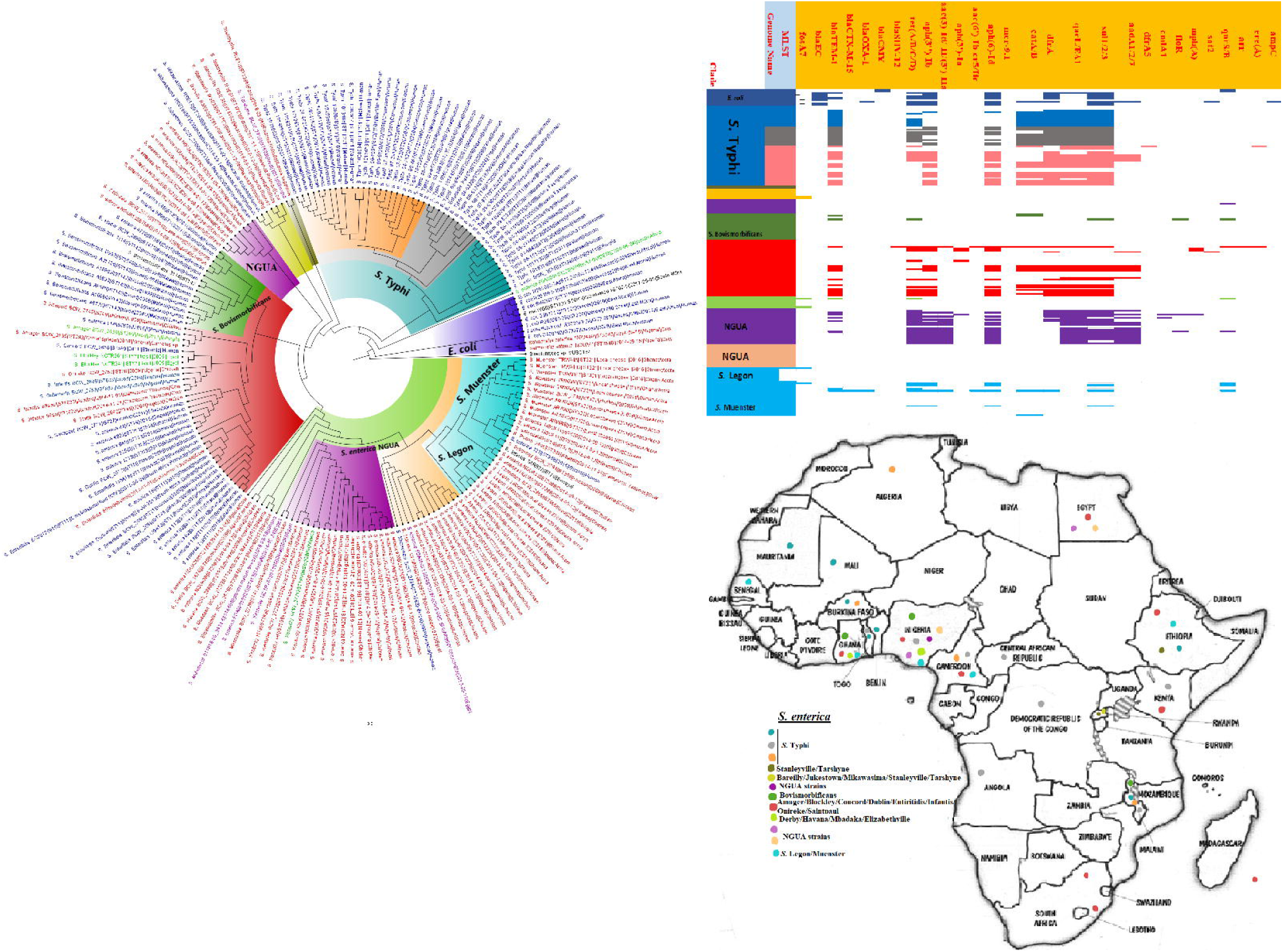
Phylogeographic distribution of *Salmonella enterica* clades and associated resistomes in Africa. *Salmonella enterica serovars* were mainly host-specific, with *S*. Typhi, *S*. Typhimurium, *S*. Entiritidis, and *S*. Bovismorbificans being isolated from humans whilst serovars such as *S*. Muenster, *S*. Legon, NGUA, *S*. Salamae, and *S*. Wilhelmsburg strains were animal-associated. In general, *S*. enterica clades were of diverse phylogeographic distribution, but clustered in Southern Africa, Madagascar, DRC, Sudan, Comoros, Cameroon, East, West and North Africa. The ARGs in *S*. enterica were serovar and clade-specific, with *S*. Typhi, *S*. Typhimurium, *S. enterica* NGUA strains hosting most ARGs such as TEM, *tet(A/B/C/D), aph(6’)-Id, aph(3”)-Ib, catA/B, dfrA, qacL/*Δ*E1*, and *sul1/2/3;* notably, *S*. Typhi clades A1, B1-B3 (Fig. S9A) mostly harboured these ARGS. Inter-country as well as human-animal-environment dissemination of isolates of the same clade were observed. Isolates from humans, animals, the environment and plants are respectively coloured as blue, red, mauve/pink and green on the phylogeny tree.

*C. coli/jejuni* strains were reported in substantial numbers from animals, which are their natural hosts ^47^, as well as from human and environmental specimens, albeit few genomes (all from South Africa, including *C. concisus* from human faeces) of these species were available from the continent (Table 1; Fig. S15). As well, they were not as geographically distributed as *S. enterica* as they were reported from only Botswana (human excreta and chicken caecum), Cameroon (household water), South Africa (human excreta and river water), and Tanzania (cattle milk/beef). Moreover, they harboured relatively fewer ARGs and had generally lower resistance rates, albeit resistance to ampicillin, azithromycin, ciprofloxacin, erythromycin, nalidixic acid, tetracycline, was high (Tables S1-S3). Interestingly, the *C. coli/jejuni* isolates were mostly multiclonal, suggesting evolutionary versatility and polyclonal dissemination. *Campylobacter spp*. are implicated in many diarrhoeal cases and are the major cause of human bacterial gastroenteritis worldwide, causing fatal infections in infants, the elderly and immunocompromised patients ^47^. Hence, the little data available on this pathogen is disturbing as it makes it difficult to effectively plan appropriate interventions. However, their presence in humans, animals and the environment in substantial numbers shows their host adaptability, making them ideal candidates for One Health surveillance studies.

*V. cholerae*, another common food- and water-borne diarrhoea-causing pathogen implicated in recurring outbreaks in many parts of Africa ^13,48^, was also reported in substantial numbers from human and more importantly, from environmental sources in several countries (Table 1; Fig. 8 & S2). The higher isolation of *V. cholerae* ST69 and ST515 clones across several countries in Southern, Eastern, and Western African countries, which clustered within three main clades having very close evolutionary distance and highly conserved but rich resistome repertoire, shows the presence of same and highly similar strains (with little evolutionary variation) circulating in Africa and causing recurring outbreaks with high morbidities and mortalities. Just as *S*. Typhi and *Campylobacter spp*., *V. cholerae* also cause serious diarrhoea in addition to vomiting in patients and has been implicated in death in untreated patients within hours ^13,48^. Subsequently, the large ARGs diversity in these strains is concerning. Indeed, a carbapenemase gene, termed *bla*_VCC-1_, mediating resistance to carbapenems and most β-lactams, has been recently detected in *Vibrio spp*. ^16,49,50^, although this was not found in any of these isolates.

**Figure 8.**
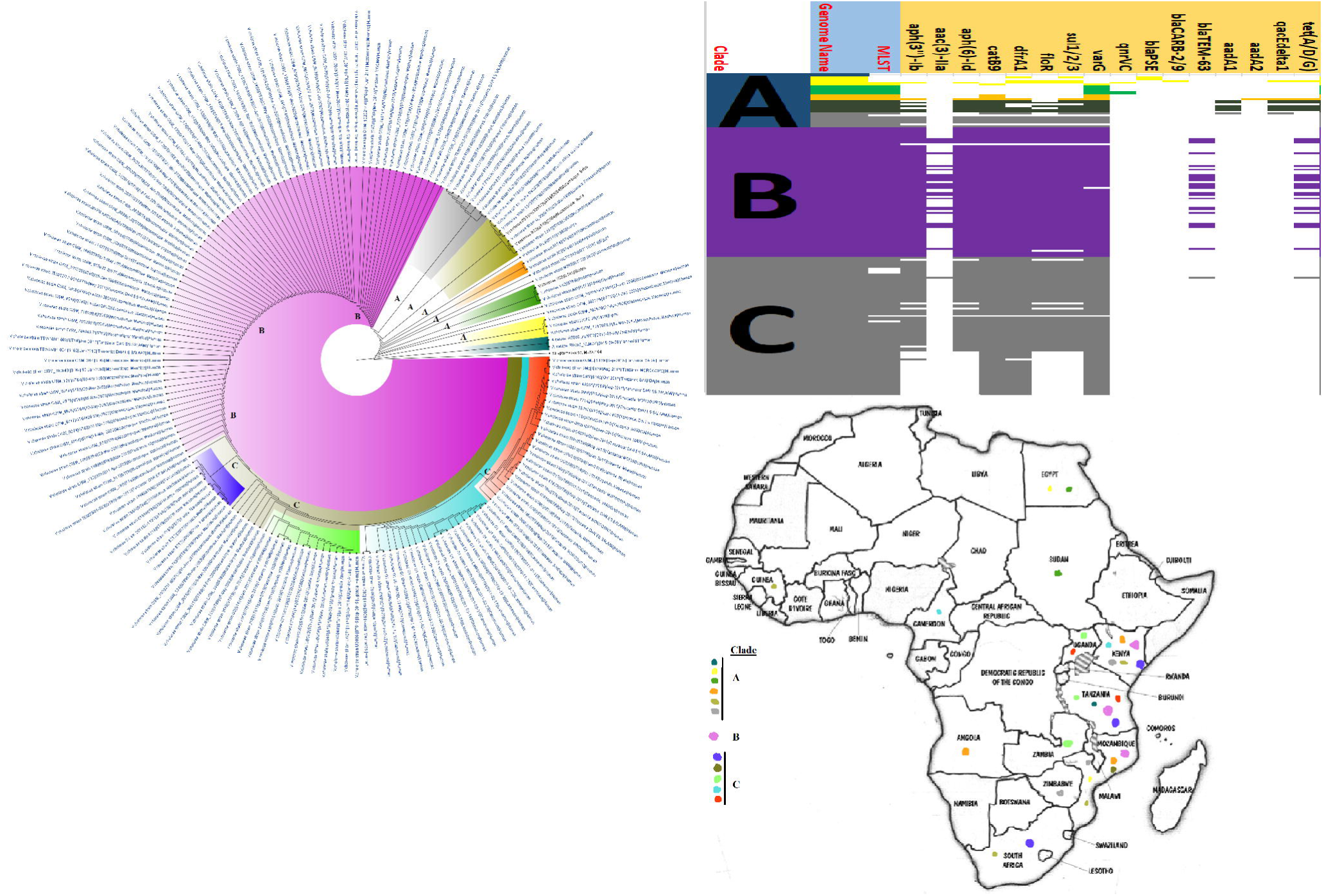
Phylogeographic distribution of *Vibrio cholerae* clades and associated resistomes in Africa. *V. cholerae* was only reported from humans in Southern and Eastern Africa, with few from Cameroon, Guinea, Sudan and Egypt. Members of clade B, which were very closely clustered together with little evolutionary variations, were mainly located in Kenya, Mozambique and Tanzania. Clades A and C were more geographically diverse with more evolutionary variations. As supported by the resistome data, clade B had almost uniform resistomes throughout, followed closely by clade C and A, which had the least repertoire of resistomes. Conserved within clades B and C were *aph(3”)-Ib, aph(6’)-Id, catB9, dfrA1, floR, sul1/2/3* and *varG*. Other ARGs in clade B were TEM-63 and *tet(A/D/G); aadA1/2 and qacL/*Δ*E1* were only found in clade A. Inter-country dissemination of isolates of the same clade were observed. Isolates from humans, animals, the environment and plants are respectively coloured as blue, red, mauve/pink and green on the phylogeny tree.

Non-fermenting Gram-negative bacilli such as *P. aeruginosa, A. baumannii, S. maltophilia* and *A. hydrophilia* are known opportunistic nosocomial pathogens with intrinsic resistance to several antibiotics ^51–55^. Particularly, *P. aeruginosa* and *A. baumannii*, which were two of the commonest pathogens with most ARGs in Africa, are commonly implicated in several difficult-to-treat and fatal clinical infections worldwide ^51–55^. Thus, the higher resistome diversity, geographical distribution, isolation and resistance rates of these pathogens are not surprising. Whereas OXA-23/51-like carbapenemases are known to be common in *A. baumannii* ^34,56^, the uniform presence of OXA-48-like carbapenemases in *P. aeruginosa* genomes from Africa is very worrying, particularly given the wider geographical distribution and ubiquity of this pathogen (Fig. 6)^51,54^. Owing to the broad β-lactams spectrum of carbapenemases and the importance of β-lactams in treating bacterial infections, the presence of these and other ARGs in these pathogens with high intrinsic resistance in Africa is a cause for concern ^4,34,56^. Given the difficulty in treating infections caused by non-fermenting Gram-negative bacilli, it is quite refreshing to note that *S. maltophilia* and *A. hydrophilia* were less isolated with little or no ARGS.

Important sexually transmitted infections such as gonorrhoea (*N. gonorrhoeae*) and non-gonococcal urethritis (*M. genitalium*), respiratory infections such as pneumonia (*M. pneumoniae*), cystic fibrosis (aggravated by *B. cepacia*) and whooping cough (*B. pertussis*), as well as cerebro-spinal infections such as meningitis (*N. meningitidis*) are caused by GNB, killing millions of people annually ^6–15,57^. Unfortunately, only *N. meningitidis* genomes were reported from Africa (17 countries), although both *N. meningitidis* (Egypt and Niger (serogroups C & W)) and *N. gonorrhoeae* (only Kenya) were found in the literature (Table S6; Fig. 5). Notably, *tet*(B) was the sole ARG found in *N. meningitidis* genomes, particularly clades B5, C and D, whereas several other mechanisms (*gyrA, penA* and *rpoB*) were reported in the literature (Table S2). These differences in the literature and genomic resistomes is quite interesting as some of the genomes were from Niger. Within each *N. meningitidis* clade were isolates from different countries, suggesting inter-boundary transmission. This is not surprising given the high transmissibility of *N. meningitidis* and the inter-country trade existing within the Sahel and West African nations with the highest concentration of this pathogen (Table S6; Fig. 5)^8^.

Worryingly, two *N. meningitidis* strains from Egypt were MDR to clinically important antibiotics such as ciprofloxacin, cefotaxime, amikacin, ampicillin, penicillin and meropenem. Sadly, reports of MDR *N. gonorrhoeae* that are highly resistant to first-line anti-gonococcal drugs such as third-generation cephalosporins and azithromycin are increasing, prompting revisions in treatment guidelines ^6,57–59^. For instance, MDR *N. gonorrhoeae* that were only treatable with carbapenems have been reported in the UK and Australia ^57–59^.

Unfortunately, *N. gonorrhoeae* resistance rates were not calculable due the absence of *N. gonorrhoeae* ASTs. Indeed, the use of molecular tests to determine *N. gonorrhoeae* resist^57^ance, although fast, is unable to provide AST data that is critical to guide treatment. The shift to molecular tests is thus making *N. gonorrhoeae* AST data scarce and could account for the dearth of information on *N. gonorrhoeae* AST in the included articles^57^.

*M. genitalium* genomes from Africa were not found, although they have been reported in the South Africa as having *gyrA* and *parC* resistance determinants (Table S2). However, *M. pneumoniae* genomes, obtained from Southern, Western, Eastern and Northern Africa, harboured no known ARGs. Other species such as *M. gallinarum, M. gallinaceum, M. pullorum, M. mycoides* and *M. capripneumoniae* were found in animals whilst *M. arginini* were isolated from the environment. Like *Neisseria spp*., increasing macrolide resistance in *Mycoplasma spp*. are being reported, making them less sensitive to azithromycin.

Although *B. cepacia* does not cause cystic fibrosis, it aggravates it ^10^. *B. cepacia* are inherently resistant to treatment by most antibiotics and they normally occur alongside *P. aeruginosa* in cystic fibrotic lungs, where they can cause persistent infections and death ^10,60^. In general, *Burkholderia spp*. were very few throughout Africa, in both literature and published genomes, with no known resistance gene being reported (Fig. S14). *B. pertussis*, the causative agent of whooping cough, a major childhood killer disease that has been killing infants for thousands of years ^14^, was rarely reported in the literature and had fewer public genomes from only Kenya and South Africa. In fact, the fewer reports of this infection in the literature does not reflect the epidemiology of whooping cough on the continent as it kills many infants in Africa yearly ^61^, suggesting that most clinical cases are not published. Although vaccinations have reduced the mortality and morbidity rates associated with this pathogen, variations in the pathogen is reducing the efficacy of the vaccine and increasing its re-emergence globally ^61^.

Whilst other Enterobacteriaceae (GNB) species such as *S. marcescens, Citrobacter spp*., *Providencia spp*., *Proteus mirabilis*, and *M. morganni* were relatively less isolated than *E. coli* and *K. pneumoniae*, they did harbour a rich and diverse collection of resistomes in different clones/clades across Africa, although many of these were mainly reported from South Africa, followed by Tanzania, Nigeria, Senegal and Egypt (Fig. S16, S18, & S21-23). These species have been implicated in fatal infections such as sepsis and wound infections ^31,34,39,62,63^, bearing critical ARGs such as ESBLs and carbapenemases (Table S2). On the other hand, rarely isolated/reported GNB species such as *Trabulsiella spp*., *Psychrobacter spp*., *Myroides spp*., *Chromobacterium spp*., *Brevundimonas spp*. and *Bacillus spp*. were mainly obtained from the environment or termites in very few countries and harboured no ARGs. As well *A. faecalis, Leclercia adecarboxylata, Pantoea spp*. and *Raoultella spp*., *which* were mainly isolated from human specimen except for *Pantoea spp*. that was also obtained from animals and humans, also contained no known ARG and were mainly restricted to few geographical areas except *Pantoea spp*. that was widely distributed across Africa (Tables 1, S1-S3 & Fig. S11, S20, S24-S25).

### Future perspectives & Conclusion

The backbone to efficient diagnosis and treatment of infections is rapid, effect, simple and cheap diagnostics and skilled laboratory scientists ^64–66^. Without appropriate diagnostics, the aetiology of many infectious diseases, the genotype/clone of the infecting organism and its resistance mechanisms cannot be known, and a proper therapeutic choice cannot be made ^64–66^. This describes the situation in many African countries, making several preventable infections emerge subtly into large scale outbreaks ^65,66^. As shown in Table 2, simpler phenotypic diagnostic tests with a longer turnaround time of at least 24 hours were more commonly used in Africa whilst complex, skill-requiring and expensive tests like whole-genome sequencing and Vitek were hardly used. These challenges affect the fight against infectious diseases and make surveillance studies on the continent difficult ^64,67^.

It is worthy of consideration that as large as the number of articles and genomes used in this study are, the dearth of genome sequencing and molecular ARG surveillance in many African countries, influenced by low funding, absence of molecular diagnostic laboratories, and inadequate skilled personnel, affect the comprehensiveness of the phylogeography and resistome evolutionary epidemiology ^4^. This is particularly true for certain species such as *V. cholerae, C. coli/jejuni, N. meningitidis/gonorrhoeae, Mycoplasma genitalium, Providencia spp*., *M. morganii, C. freundii, P. mirabilis, B. cepacia*, and *B. pertussis*, which are very important clinical pathogens implicated in substantial morbidities and mortalities ^19^.

In summary, MDR GNB clinical pathogens implicated in high morbidities and mortalities are circulating in Africa in single and multiple clones, shuttling diverse resistomes on plasmids, integrons, insertion sequences and transposons from animals, foods, plants and the environment to humans. A comprehensive One Health molecular surveillance is needed to map the transmission routes and understand the resistance mechanism of these pathogens to inform appropriate epidemiological interventions.

## Data Availability

All source data used for this article are included in the supplementary Tables and Files.

## Author contributions

JOS conceived and designed the study, searched the literature, analysed the data and undertook all bio-informatic analyses, image designs and tabulations and wrote the paper. MAR searched the literature, extracted the data and resistance genes from NCBI into Excel sheets and undertook frequency analyses of the raw data.

## Transparency declaration

The authors declare no conflict of interest

## Acknowledgements

We are exceptionally grateful to Dora Osei Sekyere of the University of Education Winneba, Kumasi Campus, Ghana, for aiding in the curation, annotation and design of the genomic and phylogenomic metadata and trees.

---

**Figure 1**. The PRISMA flow-diagram was used to summarise the literature search methodology, databases used, and the inclusion and exclusion criteria adopted in getting the final 146 manuscripts used in this qualitative and quantitative analyses. Besides the included literature, 3028 publicly available genomes were downloaded from PATRIC (https://www.patricbrc.org/)/NCBI‘s Genbank and analysed to determine their resistomes and evolutionary phylogeography.

**Figure S1**. Species distribution and frequencies per animal, human and environmental sources in respective Africa countries. The major pathogens of medical importance found in Africa included *E. coli, K. pneumoniae, S. enterica, P. aeruginosa, A. baumannii, Campylobacter coli/jejuni, Proteus mirabilis, Enterobacter spp*., *N. gonorrhoea/meningitidis* and *V. cholerae*, and these were mainly found in humans, followed by animals and the environment. These were mainly reported from Algeria, Burkina Faso, Egypt, Ghana, Kenya, Libya, South Africa, Tanzania and Tunisia in humans. South Africa, Tanzania and Nigeria, reported the highest concentrations of environmental species. Notably, *E. coli* and *S. enterica*, and to a lesser extent *Campylobacter coli/jejuni, Klebsiella spp*., and *Pseudomonas spp*., were the most isolated species from animals in the reporting countries

**Figure S2**. Frequency distribution of Gram-negative bacterial species isolated from human, animal and environmental specimens and their associated antibiotic resistance genes (ARGs) and mobile genetic elements (MGEs). *E. coli* was the most isolated species from animals, humans and the environment. There were more species from human sources than animal and environmental specimens, with *Klebsiella pneumoniae, Salmonella enterica, Pseudomonas aeruginosa* and *Acinetobacter baumannii* being also common in human samples. CTX-M, TEM, OXA, SHV, NDM, AAC(6’), AAD and QNR enzymes were common resistance mechanisms in clinical isolates whilst TET, SUL, TEM, CTX-M and CMY were common resistance mechanisms in animal isolates; TET and TEM resistance mechanisms were more isolated in environmental samples. There were more MGEs characterisation in human isolates than animal and environmental strains, with Class 1 integrons and IncF/IncX plasmids being very common in human isolates.

**Figure S3**. Clonal distribution frequency of Gram-negative bacterial species isolated from human, animal and environmental specimens. The clonality or genotypes of *E. coli, K. pneumoniae, A. baumannii, Campylobacter coli/jejuni, V. cholerae, S. marcescens, N. gonorrheae*/*meningitidis, S. enterica* Enteritidis/Typhimurium were the main data reported by the included countries. Common clones such as *E. coli* ST131, ST38, ST410, *E. coli* group A/B/D (in Egypt and Algeria) were found in humans and to some extent, in animals and the environment. Clones of *A. baumannii* (ST1, ST2, ST218, ST208, ST415, ST493, ST441, ST499 and ST600 in Egypt and ST106, ST208, ST229, ST339, ST502, ST758, ST848 and ST1552 in South Africa), *K. pneumoniae* (ST15, ST39, ST152, ST154, ST101, ST147, ST466, ST2094), *V. cholerae* ST69 (Cameroon), *P. aeruginosa* (ST235 & ST233 in Egypt and ST244 & ST640 in Tanzania), *S. marcescens* types A & B (Egypt), *N. gonnorhoea* (ST1599, ST1903, ST1893 & ST11366 in Kenya), *N. meningitidis* (ST11 & ST10217 in Niger), *S*. Typhimurium ST313 and *S*. Enteritidis ST11 (Kenya) and *S*. Typhi ST1 (Zambia) were very common in humans.

**Figure S4**. ARGs reported from animals, humans and environmental Gram-negative bacterial isolates from the included articles per country. The frequency distribution of the various ARGs that were isolated from the Gram-negative bacterial species shows that CTX-M, TEM, SHV, OXA, QnrA/B/D/S, AAC(6’)-Ib-cr, AAC(3’)-IIa, SUL/DFRA enzymes were common in selected countries/regions. In all, mechanisms mediating resistance to β-lactamases were common in all countries followed by fluoroquinolones, sulphamethoxazole-trimethoprim, aminoglyclosides, tetracycline, chloramphenicol, and erythromycin. Tetracycline resistance mechanisms were more common in animal isolates than human and environmental isolates; however, sulphamethoxazole-trimethoprim resistance genes were more common than those of tetracycline (*tet* genes).

**Figure S5**. Mobile genetic elements (MGEs) found in species isolated from animals, humans, and the environment in Africa.

**Figure S6**. Mobile genetic elements (MGEs) and associated antibiotic resistance genes (ARGs) found in species isolated from animals, humans, and the environment.

**Figure S7**. Phylogeographic distribution of *Escherichia coli* clades and associated resistomes in Africa (A & B). The *E. coli* clades were mostly from humans, mainly distributed in West and East Africa, Egypt and South Africa. Relatively few were from the environment and animals. The ARGs (*tet (A/B/C/D), bla*_TEM-1_, *sul, aph(3”)-Ib, aph(6)-Id*, and *dfrA*) were mostly conserved across the various clades, which were not region-specific, but mixed up. Inter-country as well as human-animal-environment dissemination of isolates of the same clade were observed. Isolates from humans, animals, the environment and plants are respectively coloured as blue, red, mauve/pink and green on the phylogeny tree.

**Figure S8**. Phylogeographic distribution of *Klebsiella pneumoniae* clades and associated resistomes in Africa. *Klebsiella pneumoniae* was mainly from humans, with a few being isolated from plants and animals. The various clades were mixed up in South Africa, Tanzania, Uganda, West and North Africa. Strains from humans shared very close phyletic relationship with strains from animals and plants. The clades harboured many conserved ARGs (n=14): *aac(3’)-Ia/IIa, aac(6’)-IIa/Ib-cr, aph(3”)-Ib, aph(6’)-Id, bla*_CTX-M_, *bla*_OXA_, *bla*_SHV_, *bla*_TEM_, *aadA, catA/B, dfrA, fosA, oqxAB, sul1/2*. Other ARGs that were substantially found in *K. pneumoniae* included *arr, mph(A/E), qnrA/B/S, qacE*Δ*1* and *bla*_NDM-1/5._ A few genes were restricted to certain clades. Inter-country as well as human-food-environment dissemination of isolates of the same clade were observed. Isolates from humans, animals, the environment and plants are respectively coloured as blue, red, mauve/pink and green on the phylogeny tree.

**Figure S9**. Phylogeographic distribution of *Salmonella enterica* clades and associated resistomes in Africa (A & B). *Salmonella enterica serovars* were mainly host-specific, with *S*. Typhi, *S*. Typhimurium, *S*. Entiritidis, and *S*. Bovismorbificans being isolated from humans whilst serovars such as *S*. Muenster, *S*. Legon, NGUA, *S*. Salamae, and *S*. Wilhelmsburg strains were animal-associated. In general, *S*. enterica clades were of diverse phylogeographic distribution, but clustered in Southern Africa, Madagascar, DRC, Sudan, Comoros, Cameroon, East, West and North Africa. The ARGs in *S*. enterica were serovar and clade-specific, with *S*. Typhi, *S*. Typhimurium, *S. enterica* NGUA strains hosting most ARGs such as TEM, *tet(A/B/C/D), aph(6’)-Id, aph(3”)-Ib, catA/B, dfrA, qacL/*Δ*E1*, and *sul1/2/3;* notably, *S*. Typhi clades A1, B1-B3 (Fig. S9A) mostly harboured these ARGS. Inter-country dissemination of isolates of the same clade were observed. Isolates from humans, animals, the environment and plants are respectively coloured as blue, red, mauve/pink and green on the phylogeny tree.

**Figure S10**. Phylogeographic distribution of *Aeromonas spp*. clades and associated resistomes in Africa. No known ARGs could be found in the genomes, which were all from South African environmental specimens. Isolates from humans, animals, the environment and plants are respectively coloured as blue, red, mauve/pink and green on the phylogeny tree.

**Figure S11**. Phylogeographic distribution of *Bacillus spp*. clades and associated resistomes in Africa. There were five main clusters, comprising various species of this genus, which were from animals (e.g. *B. anthracis, B. cereus, B. thuringensis*), humans (*B. anthracis*) environment (e.g. *B. anthracis, B. cereus, B. thuringensis*) and plants (e.g. *B. subtilis, B. halotolerans, B. velenzensis*) in Southern Africa, DRC, Kenya, Cameroon, B. Faso, Senegal, and North Africa. No known ARGs could be found in the genomes. Isolates from humans, animals, the environment and plants are respectively coloured as blue, red, mauve/pink and green on the phylogeny tree.

**Figure 12**. Phylogeographic distribution of *Bordetella spp*. clades and associated resistomes in Africa. The genomes were obtained from only Kenya from humans (*B. pertussis*) or the environment (other species). No known ARGs could be found in the genomes. Isolates from humans, animals, the environment and plants are respectively coloured as blue, red, mauve/pink and green on the phylogeny tree.

**Figure S13**. Phylogeographic distribution of *Brevundimonas spp*. clades and associated resistomes in Africa. Only a single strain from South Africa was reported throughout Africa. No known ARGs could be found in the genomes. Isolates from humans, animals, the environment and plants are respectively coloured as blue, red, mauve/pink and green on the phylogeny tree.

**Figure S14**. Phylogeographic distribution of *Burkholderia spp*. clades and associated resistomes in Africa. *Burkholderia spp*. genomes in Africa were solely from South Africa and B. Faso in plants and environmental samples. *B. cepacia* genomes were not found in Africa and no known ARGs could be found in the included genomes. Isolates from humans, animals, the environment and plants are respectively coloured as blue, red, mauve/pink and green on the phylogeny tree.

**Figure S15**. Phylogeographic distribution of *Campylobacter spp*. clades and associated resistomes in Africa. The included genomes were all from human (*C. concisus/jejuni*) and animal (*C. fetus*) specimens from South Africa with no known ARGs in the genomes. Isolates from humans, animals, the environment and plants are respectively coloured as blue, red, mauve/pink and green on the phylogeny tree.

**Figure S16**. Phylogeographic distribution of *Citrobacter spp*. clades and associated resistomes in Africa. The *Citrobacter spp*. genomes were from humans and water in South Africa, Tanzania, Nigeria, and Senegal. *bla*_CMY_ was the only ARG conserved in all the clades whilst clade I had more ARGs; generally, the ARGs were not clade specific, but isolate-specific. Isolates from humans, animals, the environment and plants are respectively coloured as blue, red, mauve/pink and green on the phylogeny tree.

**Figure S17**. Phylogeographic distribution of *Enterobacter spp*. clades and associated resistomes in Africa. *E. aerogenes, E. asburiae, E. cloacae, E. hormaechei*, and *E. xiangfangensis* were obtained from animals, humans, environment (water) and plants in mainly South Africa, Zambia, West and North Africa. The *Enterobacter spp*. had a rich resistome repertoire (n≥25), comprising of ARGs such as *aac(3’)-IIa, aac(6’)-IIa/Ib3/-IIc/Ib-cr, aph(3”)-Ib, aph(3’)-I/Ia, aph(6’)-Id, bla*_ACT_, *bla*_CMH_(clade A only), *bla*_NDM,_ *bla*_CTX-M_, *bla*_OXA_, *bla*_OXA-48/181_, *bla*_SHV_, *bla*_TEM_, *aadA, catA/B, dfrA, fosA, oqxAB, sul1/2, qnrB/S, tet(A/D), mcr-1/9, arr*, and *rmtC*. Inter-country as well as human-animal-water-food dissemination of isolates of the same clade were observed. Isolates from humans, animals, the environment and plants are respectively coloured as blue, red, mauve/pink and green on the phylogeny tree.

**Figure S18**. Phylogeographic distribution of *Morganella morgannii* clades and associated resistomes in Africa. Only two isolates from humans in South Africa were obtained for all of Africa; one of these two strains had a relatively rich resistome. Isolates from humans, animals, the environment and plants are respectively coloured as blue, red, mauve/pink and green on the phylogeny tree.

**Figure S19**. Phylogeographic distribution of *Mycoplasma spp*. clades and associated resistomes in Africa. These strains were host-specific, with *M. gallinarum/pullorum/gallinaceum/mycoides/capripneumoniae* being found in animals and *M. pneumoniae* being found in humans; *M. arginini* was found in the environment. These strains were mainly found in Egypt, Cameroon, Nigeria, Tunisia, Southern and Eastern Africa. No known ARGs could be found in the genomes. Isolates from humans, animals, the environment and plants are respectively coloured as blue, red, mauve/pink and green on the phylogeny tree.

**Figure S20**. Phylogeographic distribution of *Pantoea spp*. clades and associated resistomes in Africa. *Pantoea spp*. were mainly found in the environment and in termites in South Africa, Tanzania, Rwanda, Uganda, West Africa, Algeria and Tunisia. No known ARGs could be found in the genomes. Inter-country as well as plant-animal (termite) dissemination of isolates of the same clade were observed. Isolates from humans, animals, the environment and plants are respectively coloured as blue, red, mauve/pink and green on the phylogeny tree.

**Figure S21**. Phylogeographic distribution of *Proteus mirabilis* clades and associated resistomes in Africa. *P. mirabilis* genomes were only found in human samples from Egypt and South Africa, with substantial differences in their resistomes. Isolates from humans, animals, the environment and plants are respectively coloured as blue, red, mauve/pink and green on the phylogeny tree.

**Figure S22**. Phylogeographic distribution of *Providencia rettgerii* clades and associated resistomes in Africa. All the *P. rettgerii* strains were from South Africa and contained several ARGs. Isolates from humans, animals, the environment and plants are respectively coloured as blue, red, mauve/pink and green on the phylogeny tree.

**Figure S23**. Phylogeographic distribution of *Serratia marcescens* clades and associated resistomes in Africa. The *S. marcescens* strains were from mostly humans, with one being from a nematode; they were all isolated from South Africa. NDM-1 was common in the resistomes of all the isolates, with CTX-M-3, TEM and OXA-1 ESBLs being very common in many of the strains. Isolates from humans, animals, the environment and plants are respectively coloured as blue, red, mauve/pink and green on the phylogeny tree.

**Figure S24**. Phylogeographic distribution of *Stenotrophomonas maltophilia/rhizophila*. clades and associated resistomes in Africa. These two species were found in humans, water/environment and plants (*S. rhizophila* omly) in South Africa, DRC, Nigeria, Algeria and Morocco, with no known ARGs in the genomes. Isolates from humans, animals, the environment and plants are respectively coloured as blue, red, mauve/pink and green on the phylogeny tree.

**Figure S25**. Phylogeographic distribution of *Trabusiella spp*. clades and associated resistomes in Africa. These genomes were mainly isolated from termites in South Africa, with o known ARGs found in the genomes. Isolates from humans, animals, the environment and plants are respectively coloured as blue, red, mauve/pink and green on the phylogeny tree.

**Supplementary Table S1**. An Excel Table showing the general extracted meta-data from the included articles for animal samples. This sheet contains all the information on bacterial species, clones, resistance mechanisms and associated mobile genetic elements (MGEs), and resistance rate per antibiotic for each species. Resistance rates above 50% are shown in red for easy identification by the reader. Subsequent downstream qualitative and quantitative analyses were undertaken from these Tables.

**Supplementary Table S2**. An Excel Table showing the general extracted meta-data from the included articles for human samples. This sheet contains all the information on bacterial species, clones, resistance mechanisms and associated mobile genetic elements (MGEs), and resistance rate per antibiotic for each species. Resistance rates above 50% are shown in red for easy identification by the reader. Subsequent downstream qualitative and quantitative analyses were undertaken from these Tables.

**Supplementary Table S3**. An Excel Table showing the general extracted meta-data from the included articles for environmental samples. This sheet contains all the information on bacterial species, clones, resistance mechanisms and associated mobile genetic elements (MGEs), and resistance rate per antibiotic for each species. Resistance rates above 50% are shown in red for easy identification by the reader. Subsequent downstream qualitative and quantitative analyses were undertaken from these Tables.

**Supplementary Table S4**. An Excel Table showing the metadata of all strains used for the genomic, resistome and phylogenetic analyses. Each species is placed on a single sheet within the file and contains all genomic and phenotypic information on the strains.

**Supplementary Table S5**. Pictorial representation of the resistomes of all strains per species. The resistomes of each strain per species is found within separate sheets. The clades and subclades of each species is coloured differently to show similarities and evolutionary dynamics across countries. Members of the same clade are coloured with the same colours irrespective of their MLST.

**Supplementary Table S6**. Species by species breakdown of the resistomes of each included genome. The resistomes and associated genomic and demographic strain data are shown in this file without colour codings.

## Footnotes

1 Amplified fragment length polymorphism (AFLP)

2 Multi-locus sequence typing (MLST)/core-genome MLST (cgMLST)

3 Pulsed-field gel electrophoresis (PFGE)

